# Outbreak of Covid-19 worldwide is on the decline -----Recurrent Neural Reinforcement Learning and Health Interventions to Curb the Spread of Covid-19 in the world

**DOI:** 10.1101/2020.07.08.20149146

**Authors:** Qiyang Ge, Zixin Hu, Kai Zhang, Shudi Li, Wei Lin, Li Jin, Momiao Xiong

## Abstract

As the Covid-19 pandemic soars around the world, there is urgent need to forecast the expected number of cases worldwide and the length of the pandemic before receding and implement public health interventions for significantly stopping the spread of Covid-19. Widely used statistical and computer methods for modeling and forecasting the trajectory of Covid-19 are epidemiological models. Although these epidemiological models are useful for estimating the dynamics of transmission of epidemics, their prediction accuracies are quite low. Alternative to the epidemiological models, the reinforcement learning (RL) and causal inference emerge as a powerful tool to select optimal interventions for worldwide containment of Covid-19. Therefore, we formulated real-time forecasting and evaluation of multiple public health intervention problems into off-policy evaluation (OPE) and counterfactual outcome forecasting problems and integrated RL and recurrent neural network (RNN) for exploring public health intervention strategies to slow down the spread of Covid-19 worldwide, given the historical data that may have been generated by different public health intervention policies. We applied the developed methods to real data collected from January 22, 2020 to July 30, 2020 for real-time forecasting the confirmed cases of Covid-19 across the world. We observed that the number of new cases of Covid-19 worldwide reached a peak (407,205) on July 24, 2020 and forecasted that the number of laboratory-confirmed cumulative cases of Covid-19 will pass 20 million as of August 22, 2020. The results showed that outbreak of Covid-19 worldwide has peaked and is on the decline

## Introduction

As of August 4, 2020, global confirmed cases of Covid-19 passed 18,263,542 cases, including 693, 726 deaths and has spread to 213 countries and Territories, causing an immense public health crisis. The government officers and people around the world have implemented various nonpharmaceutical interventions to slow the spread of Covid-19 [1]. These public health interventions include cessation of public gatherings, traffic restriction, stay-at-home orders, closures of schools and nonessential businesses, face mask ordinances, maintaining social distancing, quarantine, isolation and expanding virus testing. However, implementing public health interventions will cause substantial economic losses and social damage. Now the critical question is how to reopen the economy, while containing the Covid-19 pandemic. A key to correctly answering this question is to reconstruct the complex epidemic dynamic systems from the data, precisely predict the extent or duration of Covid-19, and develop algorithms to evaluate the effects of public health intervention on the transmission dynamics of Covid-19 and devise practical implementable public health interventions to control the spread of Covid-19 in the world.

Widely used statistical and computer methods for modeling of Covid-19 simulate the transmission dynamics of epidemics to understand their underlying mechanisms, forecast the trajectory of epidemics, and assess the potential impact of a number of public health measures on curbing the spread speed of Covid-19 [2–8]. Covid-19 Forecast Hub collected 48 models for Covid-19 forecasts [9]. The majority of these models are epidemiological models. Although these epidemiological models are useful for estimating the dynamics of transmission, they have some critical limitations [10,11]. First, most epidemiological models assume that the reproduction number *R* is constant. However, in the real world, the reproduction number *R* is affected by various interventions such as lockdown of the epidemic areas, travel restrictions, population mobility, social distancing, and climate factors [12]. Therefore, the reproduction number R often changes over time. The assumptions that the parameters in the model are constant will dramatically limit our ability to simulate interventions and improve prediction accuracy. Second, the epidemiological models consist of ordinary differential equations that have many unknown parameters and depend on many assumptions. Most analyses used hypothesized parameters, which often lead to poorly fitting data. Third, the successful application of public health intervention planning highly depends on the model parameter identifiability. However, some researchers show that the parameters in the complex compartmental dynamic models are unidentifiable [13]. The values of parameters cannot be uniquely determined from the real data [14]. The variances of the estimators of these parameters are very high. Fourth, the intervention measures are not explicitly included in the epidemiological models. These models lack the mechanisms to evaluate the actual effects of public health interventions on infection rates in the ongoing Covid-19 [2].

An essential issue for overcoming these limitations is to explicitly incorporate counterfactual evaluation mechanisms into the models. Reinforcement learning (RL) and counterfactual outcome can be used as a general framework for evaluating the dynamic response of Covid-19 to the intervention measures and optimizing the intervention strategy [15–22]. RL is learning actions or interventions. It arises from solving optimal control problems of partially observed Markov Decision Processes by learning an intervention policy [23].

The control problem consists of identifying the dynamic systems and optimal control design. We can view the transmission dynamics of Covid-19 as a dynamic system or Markov Decision Process. A typical dynamic system is usually modeled by nonlinear state space equations, which can in turn be transformed into recurrent neural networks (RNN) [24]. The RNN is an ideal tool to learn a partially observed Markov Decision Process. After the dynamic system or Markov Decision Process is learned from historical data, we can use RL or optimal control theory (dynamic programming for a discrete system or Pontryagin’s maximum principle for a continuous system) to infer control signal or actions, which transforms the system to the desired state [25]. RL provides a wealth of information about the consequences of actions, or information about cause and effect.

The goal of public health interventions is to contain the Covid-19 as soon as possible. However, the set of actions or health interventions for stopping the spread of Covid-19 is limited. The environments that determine the transition dynamics of Covid-19 may change rapidly over time. The future environments of Covid-19 may be substantially different from the previous one. The actions or interventions cannot be only inferred from the historical data. To fully design optimal actions or interventions in the RL may not be feasible. Therefore, we formulated the real-time forecasting and evaluating multiple public health intervention problem into off-policy evaluation (OPE) and counterfactual outcome forecasting problem within the RL framework where the aim is to estimate the response of a new public health intervention policy, given historical data that may have been generated by different public health intervention policies [26]. We interpreted the interventions as treatments where multiple interventions were implemented at different time points and the number of new cases as treatment responses. The accurate estimation of effects of public health interventions over time would allow health officers to make plans on what intervention strategies should be used and at what times to implement interventions [27].

Public health interventions including virus testing, isolation and contact tracing, travel restriction, strict self-quarantine for families, maintaining social distancing, stopping mass gatherings, closure of schools and nonessential business and vacating hotels. To quantify comprehensive intervention strategies, an intervention variable that comprehensively and abstractly measures virus testing, mobility activities and social distancing was used as an action variable in the RL.

Recurrent neural reinforcement learning (RNRL) is taken as a general framework for investigating how Covid-19 evolves under different interventions, how individual nations respond to the interventions over time, and what are optimal timings for implementing interventions. Therefore, the RNRL will provide new tools to forecast the trajectory of Covid-19 under interventions and improve public health planning and decision making.

The RNRL was applied to the surveillance data of lab confirmed Covid-19 cases in the world up to July 30, 2020. Data on the number of confirmed and new cases of Covid-19 from January 22, 2020 to July 30, 2020 were obtained from the John Hopkins Coronavirus Resource Center (https://coronavirus.jhu.edu/MAP.HTML).

## Methods

### RNRL as a framework for modeling and evaluating the effect of the interventions on the spread of Covid-19

Markov Decision Process (MDP) is a theoretic process for the RL. RL has three components: state, action and reward and consists of system identification and optimal control of design [28]. The RNRL combines the RL with RNN [23]. The RL can be viewed as an open dynamic system with a correspondent reward function (or loss function). The dynamic system can be a discrete time or continuous time dynamic system. Here we focus on discrete time dynamic systems and partially observed MDP.

Let *h_t_* ∈ *R^m^* be a hidden state, *y_t_* be the observed variable (the number of new cases), *A_t_* be an intervention variable or action variable and *x_t_* be a vector of covariates at time *t*. Consider the following dynamic system underlying the transmission dynamics of Covid-19:

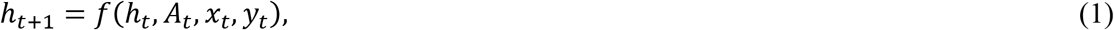

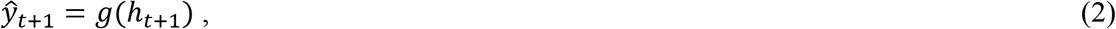

where equation (1) is the system equation, equation (2) is the observation equation, and *f,g* are two nonlinear functions. System equation (1) states that the next hidden state *h_t_*_+1_ is transitioned from the current hidden state *h_t_* and influenced by the current action or intervention *A_t_*.

The corresponding reward function is defined as *R: A* → *R*, which is a function of the current action. The reward at time *t* is defined as *R_t_ = R(A_t_*). Since the current reward may make a small contribution to the total reward in the long run, an accumulated reward over time with a possible discount factor *γ* ∈ [0,1] is defined as

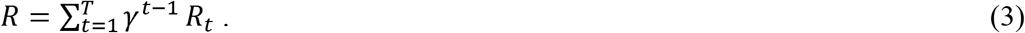

The MDP and agent (learner) generate a sequence: *h*_0_, *A*_0_, *R*_1_, *h*_1_, *A*_1_, *R*_2_,…. The RL consists of two step learning: (1) system identification and (2) optimal intervention policy learning. The reward functions in two step learning are different.

### Reward function for system identification

The system identification serves two purposes. First, since the dynamics of Covid-19 is partially observed, the hidden states should be estimated from the historical data. Second, to learn the optimal control (intervention) policy, we need to identify the system underlying the dynamics of Covid-19. It serves as a basis for the second step, optimal intervention policy learning. For the convenience of discussion, equation (2) is modified to

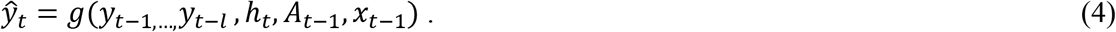

Our goal is to minimize the reward (loss) function:

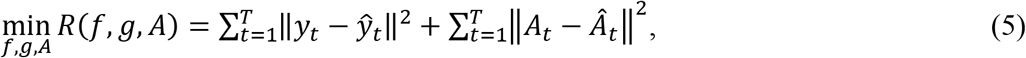

where *A* = [*A*_0_*, A*_1_,…, *A_T_*_−1_]*^T^* are estimated from the data, *f, g* functions are implemented by RNN (See Supplementary Note A)

### Reward function for optimal intervention policy learning

Inferring the optimal intervention (control) policy depends on the model identified in the previous step. In the second step, we search an optimal intervention (control) policy that minimizes the number of cumulated cases or the number of deaths. Therefore, the reward function at time *t* is defined as

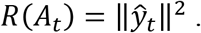

In other words, we want to make the number of new cases at time *t* as small as possible.

Let *π* be the action selection policy which determines the model’s next action *A_t_*. The action selection policy *π* which depends on the hidden state, observed data and covariates is given by

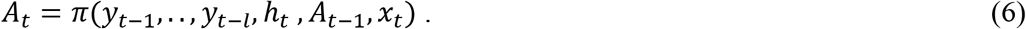

We attempt to minimize the reward function:

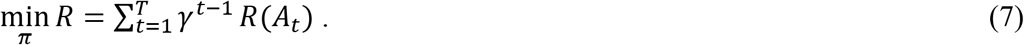

### RNN for system identification

System identification is to learn a model underlying the dynamics of Covid-19 from available historical data. The historical data includes the number of cases (new or cumulative) y_t_, the covariates *x_t_* such as age, sex, race, and the action or intervention *A_t_*. The model captures the main developments of the underlying system and explains the system evolvement beyond the observed data region. Recurrent neural networks (RNN) are a powerful tool for system identification [29]. The RNN can learn the complex dynamics within the temporal ordering of input time series of Covid-19 and use an internal memory to remember.

The RNN consists of two types of inputs and outputs: (1) internal input and output and (2) external input and output (Figure 1). The internal output of RNN can be viewed as “system state” *h_t_* which is passed to the next time step. An RNN cell receives a prior internal state *h_t−_*_1_ and a current external input: the number of cases *y_t_, …,y_t−_*_1+1_, action (intervention) *A_t_* and covariates *x_t_*, and generates a current internal state *h_t_* and an external current output 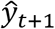 (the number of cases) at time *(t +* 1). The RNN models input the time series (past history of the number of cases of Covid-19 over time) and predict future response time series (number of cases of Covid-19 in the future with a planned sequence of interventions).

**Figure 1.**
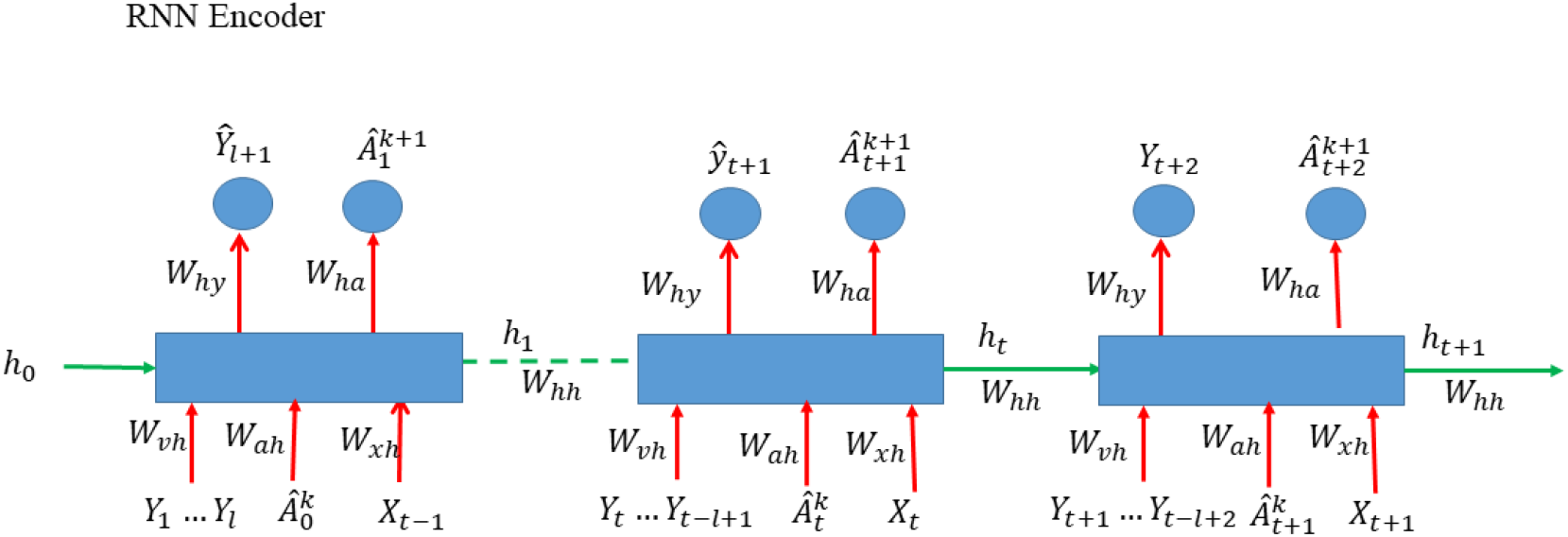
Architecture of RNN encoder.

Define the input vector *V_t_* as

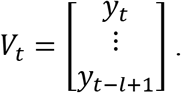

The RNN model a state transition and an output equation of the dynamic system underlying Covid-19 as follows:

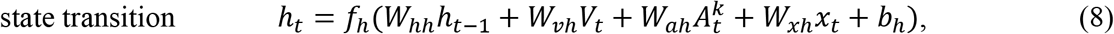

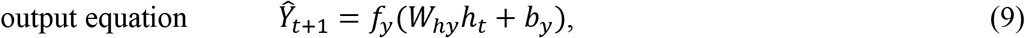

where *W_hh_* is a *m* × *m* dimensional weight matrix that connects the previous state to the current state, *W_vh_* is a *m × l* dimensional matrix, 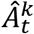 is the *k^th^* iteration of intervention measure at time *t*, *W_ah_* is a *m* dimensional vector. *W_xh_* is a *m × k* dimensional matrix, *x_t_* is a *k* dimensional vector of covariates, and 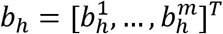 is a *m* dimensional bias vector that corrects the bias, and *f_h_* is an element-wise nonlinear activation function, and *W_hy_* is a m dimensional weight vector, *f_y_* is an activation function and *b_y_* is the bias vector of the output neurons.

In summary, using RNN to identify the system underlying the dynamics of Covid-19 can be formulated as the following optimization problem:

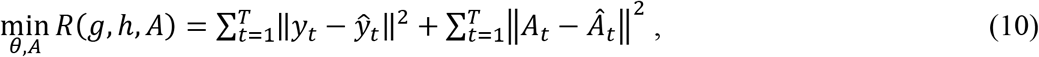

s. t.

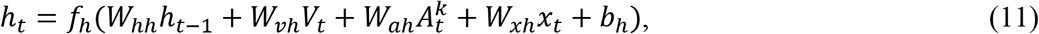

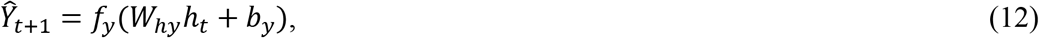

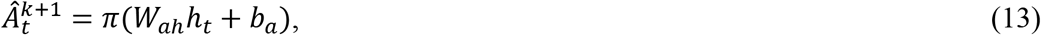

where 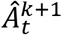 is the *(k* + 1*)^th^* iteration of intervention measure at time *t*, *π* is a nonlinear activation function, *W_ah_* is a 1×*m* dimensional matrix, and the parameters *θ* are the weight matrices and bias vectors. The above minimization problem will be solved by a backpropagation method and forward dynamic programming [27]. The detailed algorithm for training is summarized in the Supplementary Note A.

### RNN for learning actions

The main purpose of the RL is to make the best decision from historical data. The second part of the typical RL is to learn optimal control policy (Figure 2). Learning optimal control policy is usually formulated as an optimal control problem. If the state space is discrete, dynamic programming is used to find the optimal control policy [27]. If the state space is continuous, the Hamilton-Jacobi-Bellman (HJB) equation is used to solve the optimal control problem [29]. Choices of public health interventions are restricted by multiple political, cultural, technological and economic factors. Policy optimization is often practically infeasible. Therefore, we do not attempt to design optimal control actions.

**Figure 2.**
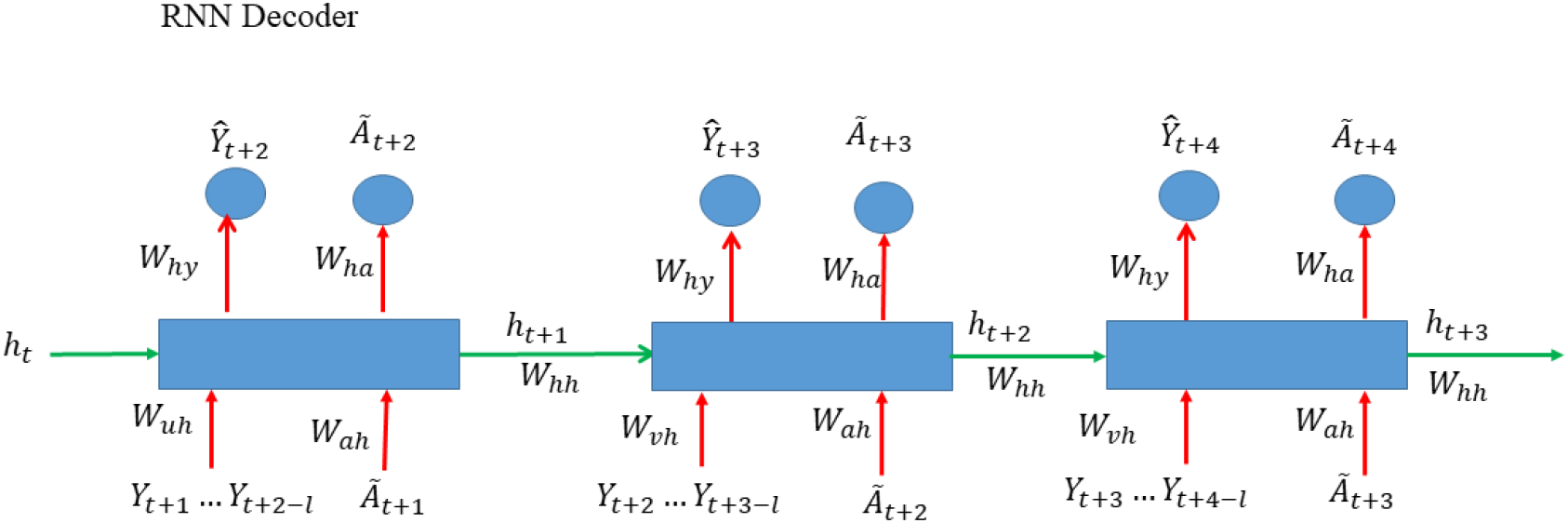
Architecture of RNN decoder.

In contrast, we use off-policy methods that evaluate or improve a policy different from that used to generate the data to select suitable actions (interventions) from a set of feasible actions (interventions). We propose to use RNN-based counterfactual action evaluation as a general framework for modeling and forecasting the spread of Covid-19 over time with multiple interventions [30]. Second RNN is used for learning counterfactual actions (interventions). The RNN forecasts the intervention response (similar to counterfactual outputs) for a given set of planned counterfactual actions (interventions) and evaluates the impact of different counterfactual actions (intervention) and their implementation times on stopping the spread of Covid-19 and provides timely selection of suitable sequence of actions (intervention) [21].

The RNN for system identification is called an encoder (Figure 1) and the RNN for action selection and evaluation is called a decoder (Figure 2). The RNN encoder models input time series (past history of the number of cases of Covid-19 over time) and predicts future response time series (number of cases of Covid-19 in the future with a planned sequence of interventions). RNN encoder was explained in the previous section. Here, we focus on the RNN decoder. Unlike the standard decoder where the decoder reconstructs back the input time series from the latent representation, the RNN decoder uses the learned features of the dynamics of Covid-19 in the RNN encoder to forecast the counterfactual response time series, given a sequence of planned counterfactual public health interventions as an input to the RNN decoder. The feature vector learned in the RNN encoder is then provided as an input to the RNN decoder which initiate prediction of the future dynamics of Covid-19 under the future counterfactual interventions (Figure 2). The RNN decoder can be represented by the following set of equations:

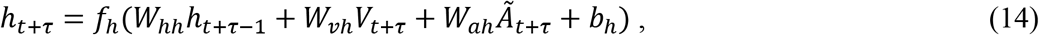

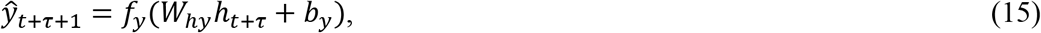

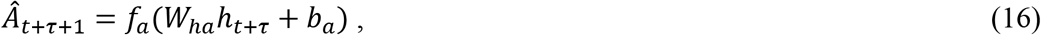

where *V_t+τ_* is defined as before, *τ ≥* 1.

The algorithm for action (intervention) evaluation and selection are summarized in Supplementary Note A.

### Data Collection

The analysis is based on surveillance data of confirmed cumulative and new Covid-19 cases worldwide as of July 30, 2020. Data on the number of cumulative and new cases and Covid-19-attributed deaths across 187 countries from January 22, 2020 to July 30, 2020 were obtained from John Hopkins Coronavirus Resource Center (https://coronavirus.jhu.edu/MAP.HTML).

### Data Pre-processing

Data were split into a training dataset (01/22-07/23, 2020) and validation dataset (07/24-07/30, 2020). All the input number of lab-confirmed cumulative cases *y_t_* was pre-processed by the following transformation: 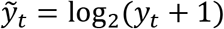. The number of new cases was calculated as 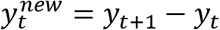.

### Mini-batches, Normalization and RNRL Flowchart

The RNRL algorithm flowchart was shown in Figure S1. We first randomly picked *k = 64* countries with *(l + τ)* length of Covid-19 time series data starting from isolated-randomly picked days to generate *k* time series with *(l + τ)* length for a mini-batch that was used for backpropagation training through time. The *l* length of time series were taken to train the RNN encoder and the *τ* length of time series were taken to train the RNN decoder. Repeat the above training processes *n* times. After the RNN encoder and decoder were trained, the trained RNN encoder and decoder were used for forecasting and evaluation. The time series *y_t−l_*_+1_*,…*, *y_t_* were fed into the trained RNN encoder, while the RNN decoder were used to forecast the time series *y_t_*_+1_*,…*, *y_t_*_+_*_τ_*. Calculate the mean value of each time series in the batch. The values of each time series were divided by their mean values.

### Forecasting Procedures

The trained RNN decoder was used to forecast the future number of new or cumulative cases of Covid-19 worldwide and for each country. The recursive multiple-step forecasting involved using a one-step model multiple times where the prediction for the preceding time step and intervention strategy were used as an input for making a prediction on the following time step. For example, for forecasting the number of new confirmed cases for the next day, the predicted number of new cases and intervention measure in one-step forecasting would be used as an observational input in order to predict the following day. Repeat the above process to obtain the two-step forecasting. The summation of the final forecasted number of new or cumulative confirmed cases for each country was taken as the prediction of the total number of new or cumulative confirmed cases of Covid-19 worldwide.

## Results

### Prediction accuracy of the dynamics of Covid-19 using RNRL

Accurate prediction of the transmission dynamics of Covid-19 is important for health decision making. To demonstrate that the RNRL was an accurate forecasting method, the RNRL was applied to the lab confirmed accumulated cases of Covid-19 across 187 countries. Figures 3 and 4 plotted reported and one-step ahead predicted time-case curves of Covid-19 in the world and top fifteen most-affected countries where blue and red curves were the number of reported and predicted cumulative cases, respectively. The top fifteen most-affected countries included US, Brazil, Russia, India, United Kingdom, Spain, Italy, Peru, Columbia, Saudi Arabia, Iran, South Africa, Chile, Mexico, and Pakistan. The average absolute of the one-step ahead prediction error in the world was 0.01596, ranging from 0.007 to 0.023. The average non-absolute and absolute of the one-step ahead prediction error in fifteen countries were −0.0172 and 0.0248, respectively. To further reliably evaluate the forecasting accuracy, we reported 7-step ahead forecasted numbers of cumulative cases and errors of Covid-19 worldwide and in 15 countries in Table 1 starting with July 24, 2020. The absolute of forecasting errors ranged from 0.00023 to 0.076.

**Figure 3.**
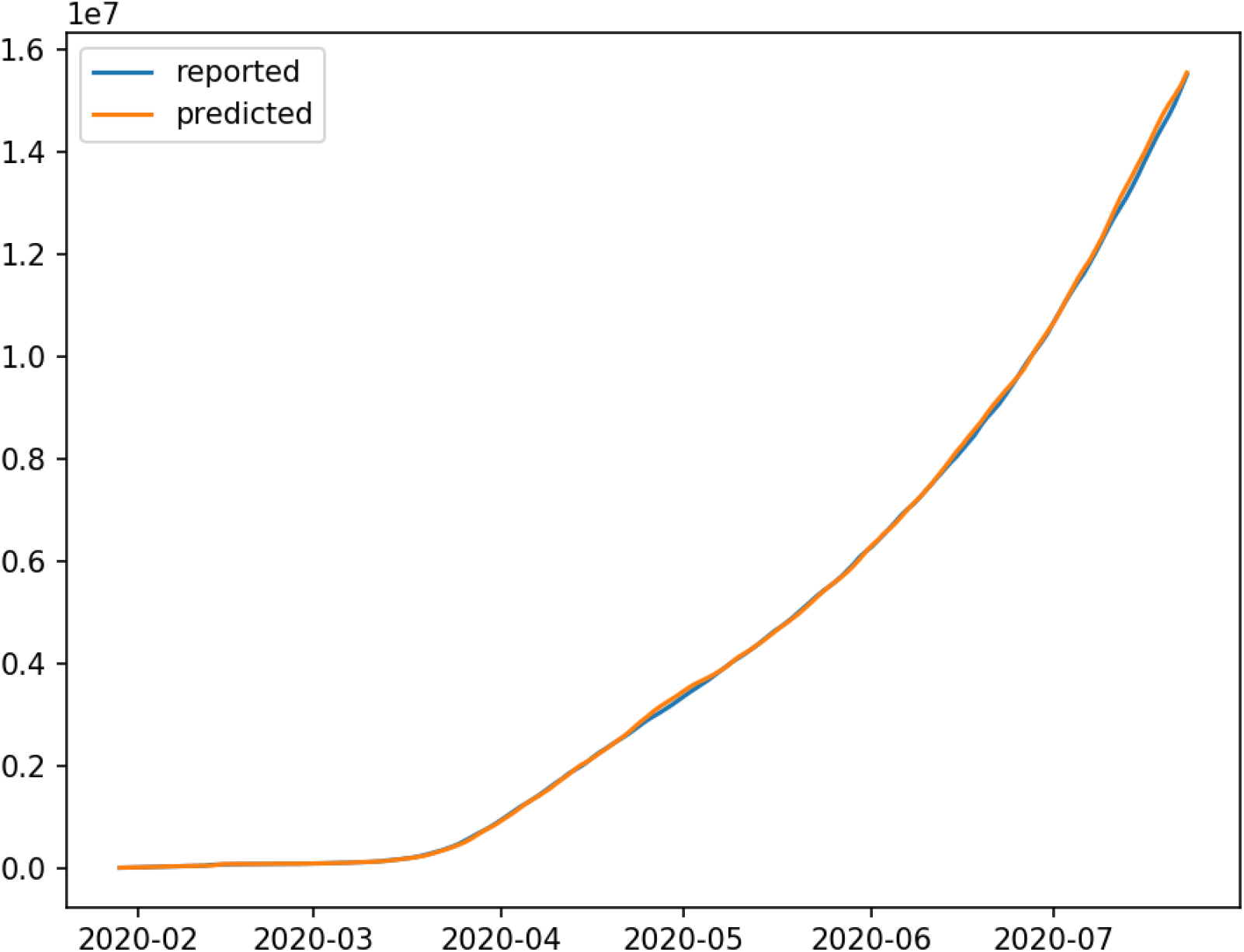
Reported and predicted time-case curves of Covid-19 worldwide where blue curve and red curve were the number of reported and predicted cumulative cases, respectively.

**Figure 4.**
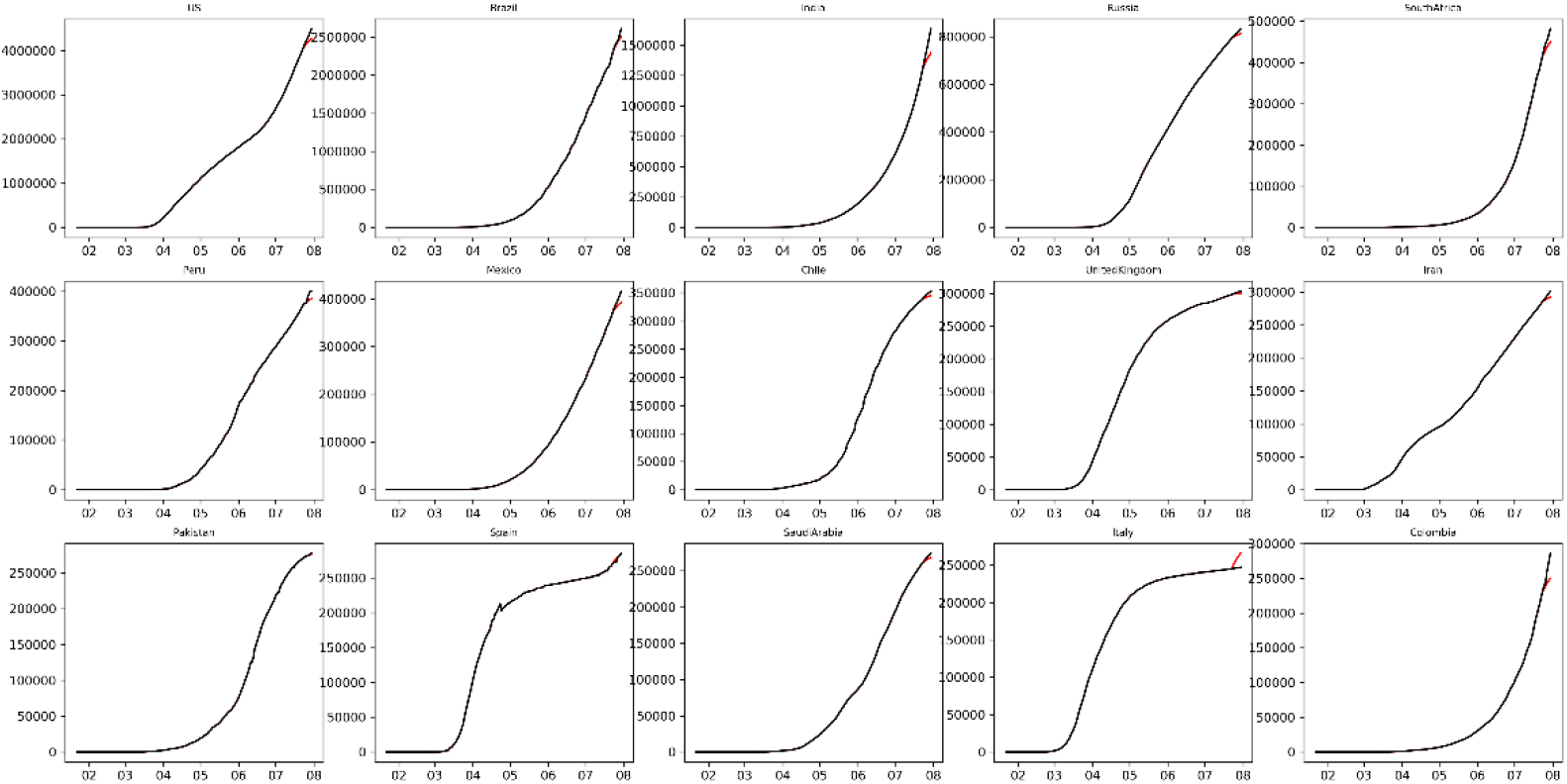
Reported and predicted time-case curves of Covid-19 in top fifteen most-affected countries where black curve and red curve were the number of reported and predicted cumulative cases, respectively.

**Table 1.** Forecasting errors of worldwide and 15 countries.

|  | Reported | Forecasted | Errors |  | Reported | Forecasted | Errors |
| --- | --- | --- | --- | --- | --- | --- | --- |
| Global |  |  |  | Chile |  |  |  |
| 7/24/2020 | 15792390 | 15902801 | 0.0070 | 7/24/2020 | 341304 | 340228 | -0.0032 |
| 7/25/2020 | 16047935 | 16215293 | 0.0104 | 7/25/2020 | 343592 | 341257 | -0.0068 |
| 7/26/2020 | 16253034 | 16546161 | 0.0180 | 7/26/2020 | 345790 | 342285 | -0.0101 |
| 7/27/2020 | 16487669 | 16859451 | 0.0225 | 7/27/2020 | 347923 | 343314 | -0.0132 |
| 7/28/2020 | 16740006 | 17098627 | 0.0214 | 7/28/2020 | 349800 | 344034 | -0.0165 |
| 7/29/2020 | 17029155 | 17333231 | 0.0179 | 7/29/2020 | 351575 | 344754 | -0.0194 |
| 7/30/2020 | 17305917 | 17556204 | 0.0145 | 7/30/2020 | 353536 | 345474 | -0.0228 |
| US |  |  |  | United Kingdom |  |  |  |
| 7/24/2020 | 4112531 | 4088453 | -0.0059 | 7/24/2020 | 299500 | 299141 | -0.0012 |
| 7/25/2020 | 4178970 | 4123199 | -0.0133 | 7/25/2020 | 300270 | 299428 | -0.0028 |
| 7/26/2020 | 4233923 | 4157945 | -0.0179 | 7/26/2020 | 301020 | 299715 | -0.0043 |
| 7/27/2020 | 4290337 | 4192691 | -0.0228 | 7/27/2020 | 301708 | 300003 | -0.0057 |
| 7/28/2020 | 4356206 | 4217013 | -0.0320 | 7/28/2020 | 302261 | 300204 | -0.0068 |
| 7/29/2020 | 4426982 | 4241335 | -0.0419 | 7/29/2020 | 303063 | 300405 | -0.0088 |
| 7/30/2020 | 4495015 | 4265657 | -0.0510 | 7/30/2020 | 303910 | 300606 | -0.0109 |
| Brazil |  |  |  | Iran |  |  |  |
| 7/24/2020 | 2343366 | 2334977 | -0.0036 | 7/24/2020 | 286523 | 285872 | -0.0023 |
| 7/25/2020 | 2394513 | 2368228 | -0.0110 | 7/25/2020 | 288839 | 287158 | -0.0058 |
| 7/26/2020 | 2419091 | 2401480 | -0.0073 | 7/26/2020 | 291172 | 288444 | -0.0094 |
| 7/27/2020 | 2442375 | 2434731 | -0.0031 | 7/27/2020 | 293606 | 289730 | -0.0132 |
| 7/28/2020 | 2483191 | 2458007 | -0.0101 | 7/28/2020 | 296273 | 290631 | -0.0190 |
| 7/29/2020 | 2552265 | 2481283 | -0.0278 | 7/29/2020 | 298909 | 291531 | -0.0247 |
| 7/30/2020 | 2610102 | 2504559 | -0.0404 | 7/30/2020 | 301530 | 292431 | -0.0302 |
| India |  |  |  | Pakistan |  |  |  |
| 7/24/2020 | 1337024 | 1320112 | -0.0126 | 7/24/2020 | 271887 | 271634 | -0.0009 |
| 7/25/2020 | 1385635 | 1342515 | -0.0311 | 7/25/2020 | 273113 | 272498 | -0.0023 |
| 7/26/2020 | 1435616 | 1364918 | -0.0492 | 7/26/2020 | 273113 | 273362 | 0.0009 |
| 7/27/2020 | 1480073 | 1387320 | -0.0627 | 7/27/2020 | 274289 | 274226 | -0.0002 |
| 7/28/2020 | 1531669 | 1403002 | -0.0840 | 7/28/2020 | 275225 | 274830 | -0.0014 |
| 7/29/2020 | 1581963 | 1418684 | -0.1032 | 7/29/2020 | 276288 | 275435 | -0.0031 |
| 7/30/2020 | 1634746 | 1434366 | -0.1226 | 7/30/2020 | 277402 | 276040 | -0.0049 |
| Russia |  |  |  | Spain |  |  |  |
| 7/24/2020 | 799499 | 797851 | -0.0021 | 7/24/2020 | 272421 | 273373 | 0.0035 |
| 7/25/2020 | 805332 | 800742 | -0.0057 | 7/25/2020 | 272421 | 275617 | 0.0117 |
| 7/26/2020 | 811073 | 803634 | -0.0092 | 7/26/2020 | 272421 | 277862 | 0.0200 |
| 7/27/2020 | 816680 | 806525 | -0.0124 | 7/27/2020 | 278782 | 280107 | 0.0048 |
| 7/28/2020 | 822060 | 808549 | -0.0164 | 7/28/2020 | 280610 | 281678 | 0.0038 |
| 7/29/2020 | 827509 | 810573 | -0.0205 | 7/29/2020 | 282641 | 283249 | 0.0022 |
| 7/30/2020 | 832993 | 812597 | -0.0245 | 7/30/2020 | 285430 | 284821 | -0.0021 |
| South Africa |  |  |  | Saudi Arabia |  |  |  |
| 7/24/2020 | 421996 | 417257 | -0.0112 | 7/24/2020 | 262772 | 262127 | -0.0025 |
| 7/25/2020 | 434200 | 423700 | -0.0242 | 7/25/2020 | 264973 | 263340 | -0.0062 |
| 7/26/2020 | 445433 | 430144 | -0.0343 | 7/26/2020 | 266941 | 264554 | -0.0089 |
| 7/27/2020 | 452529 | 436587 | -0.0352 | 7/27/2020 | 268934 | 265767 | -0.0118 |
| 7/28/2020 | 459761 | 441098 | -0.0406 | 7/28/2020 | 270831 | 266616 | -0.0156 |
| 7/29/2020 | 471123 | 445608 | -0.0542 | 7/29/2020 | 272590 | 267465 | -0.0188 |
| 7/30/2020 | 482169 | 450119 | -0.0665 | 7/30/2020 | 274219 | 268315 | -0.0215 |
| Peru |  |  |  | Italy |  |  |  |
| 7/24/2020 | 375961 | 374220 | -0.0046 | 7/24/2020 | 245590 | 249846 | 0.0173 |
| 7/25/2020 | 375961 | 376407 | 0.0012 | 7/25/2020 | 245864 | 253001 | 0.0290 |
| 7/26/2020 | 375961 | 378594 | 0.0070 | 7/26/2020 | 246118 | 256156 | 0.0408 |
| 7/27/2020 | 389717 | 380781 | -0.0229 | 7/27/2020 | 246286 | 259312 | 0.0529 |
| 7/28/2020 | 395005 | 382312 | -0.0321 | 7/28/2020 | 246488 | 261520 | 0.0610 |
| 7/29/2020 | 400683 | 383842 | -0.0420 | 7/29/2020 | 246776 | 263729 | 0.0687 |
| 7/30/2020 | 400683 | 385373 | -0.0382 | 7/30/2020 | 247158 | 265938 | 0.0760 |
| Mexico |  |  |  | Colombia |  |  |  |
| 7/24/2020 | 378285 | 375513 | -0.0073 | 7/24/2020 | 233541 | 231546 | -0.0085 |
| 7/25/2020 | 385036 | 378874 | -0.0160 | 7/25/2020 | 240795 | 235167 | -0.0234 |
| 7/26/2020 | 390516 | 382235 | -0.0212 | 7/26/2020 | 240795 | 238788 | -0.0083 |
| 7/27/2020 | 395489 | 385596 | -0.0250 | 7/27/2020 | 257101 | 242409 | -0.0571 |
| 7/28/2020 | 402697 | 387949 | -0.0366 | 7/28/2020 | 267385 | 244944 | -0.0839 |
| 7/29/2020 | 408449 | 390301 | -0.0444 | 7/29/2020 | 276055 | 247479 | -0.1035 |
| 7/30/2020 | 416179 | 392654 | -0.0565 | 7/30/2020 | 286020 | 250014 | -0.1259 |

### Transmission Dynamics of Top Fifteen Most-affected Countries

Figures 5 and 6 plotted the reported and forecasted trajectory of the new and cumulative cases of Covid-19 in the top fifteen most-affected countries, respectively. Tables S1 and S2 listed one month forecasted number of new and cumulative cases of Covid-19 in the top fifteen most-affected countries, respectively. We observed several remarkable features. First, keeping the current intervention measure, all the top 15 most-affected countries have passed the peak. Second, the spread of Covid-19 in all the top most-affected countries was curbed. The forecasted number of new cases in 12 countries on August 22, 2020 was less than 1,000 (Chile: 108, Colombia: 958, Iran: 154, Italy: 23, Mexico: 446, Pakistan: 64, Peru: 701, Russia: 314, Saudi Arabia: 109, South Africa: 632, Spain: 256, and United Kingdom: 46), the number of cases in 3 countries was less than 5,000 (Brazil: 4,183, India: 2,974, US: 3,934).

**Figure 5.**
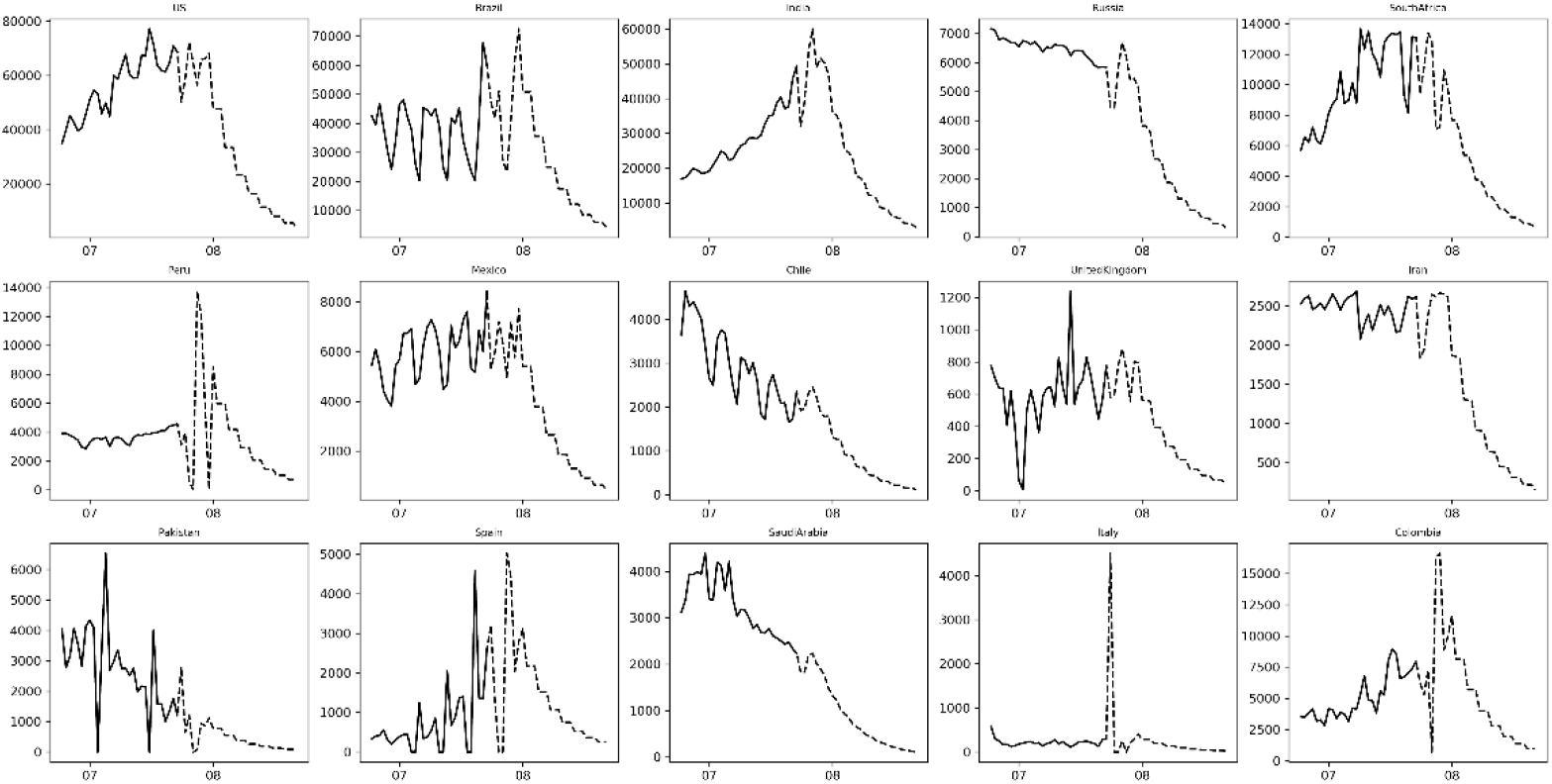
The trajectory of the new cases of Covid-19 in the top fifteen most-affected countries where solid curve and dotted curve were the number of reported and predicted new cases, respectively.

**Figure 6.**
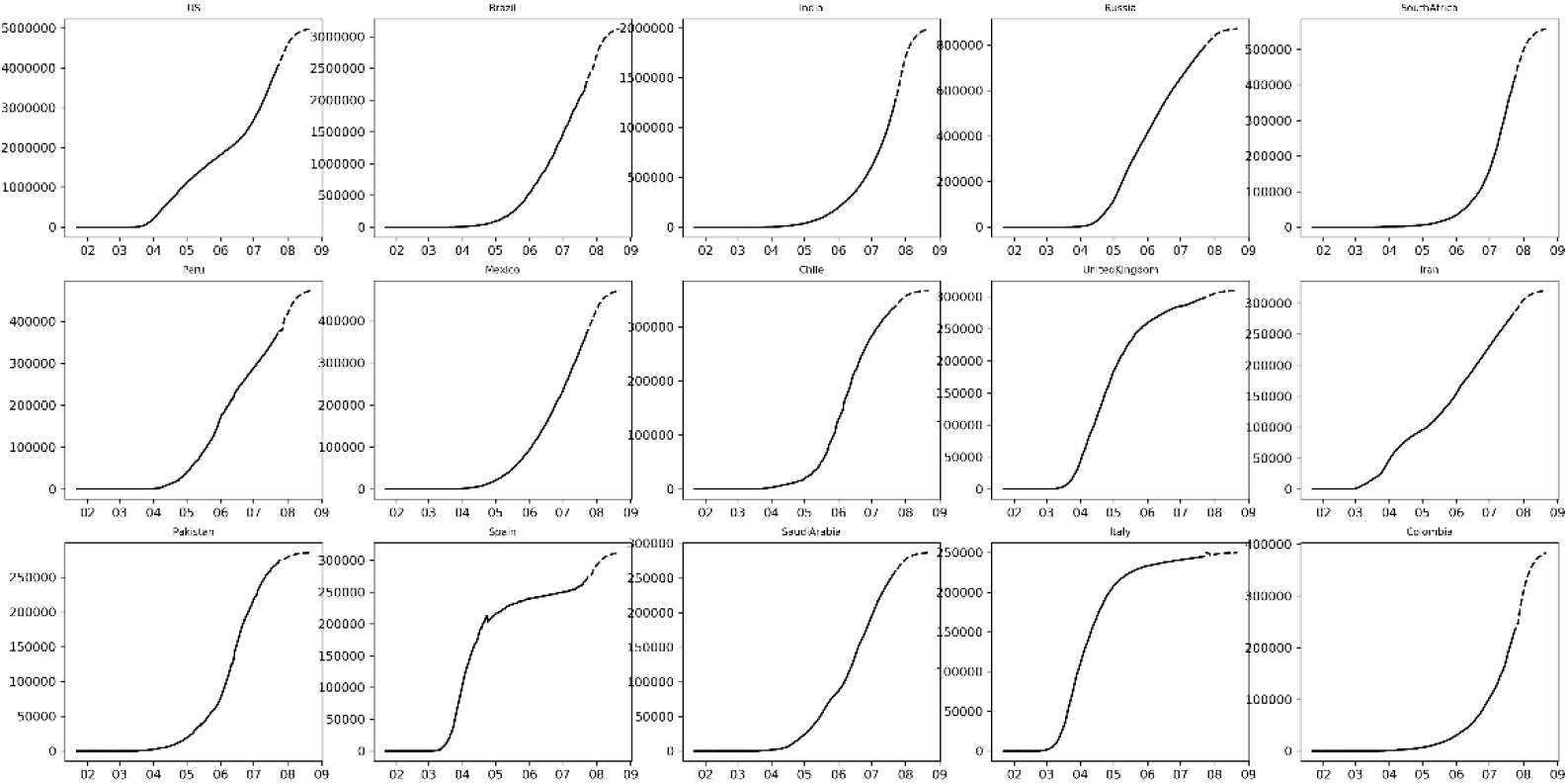
The trajectory of the cumulative cases of Covid-19 in the top fifteen most-affected countries where solid curve and dotted curve were the number of reported and predicted cumulative cases, respectively.

### Outbreak of Covid-19 worldwide has peaked and is on the decline

We observed that the number of new cases of Covid-19 worldwide reached a peak (407,205) on July 24, 2020 and forecasted that the number of new cases will reduce to 29,517 on August 22, 2020. The reported and forecasted curves of the number of new cases and the number of cumulative cases of Covid-19 worldwide were shown in Figures 7 and 8, respectively. Table 2 summarized the number of cumulative and new cases of Covid-19 worldwide, starting from July 24, 2020 to August 22, 2020. The figures and table showed the forecasted number of new cases and cumulative cases of Covid-19 for 30 days.

**Figure 7.**
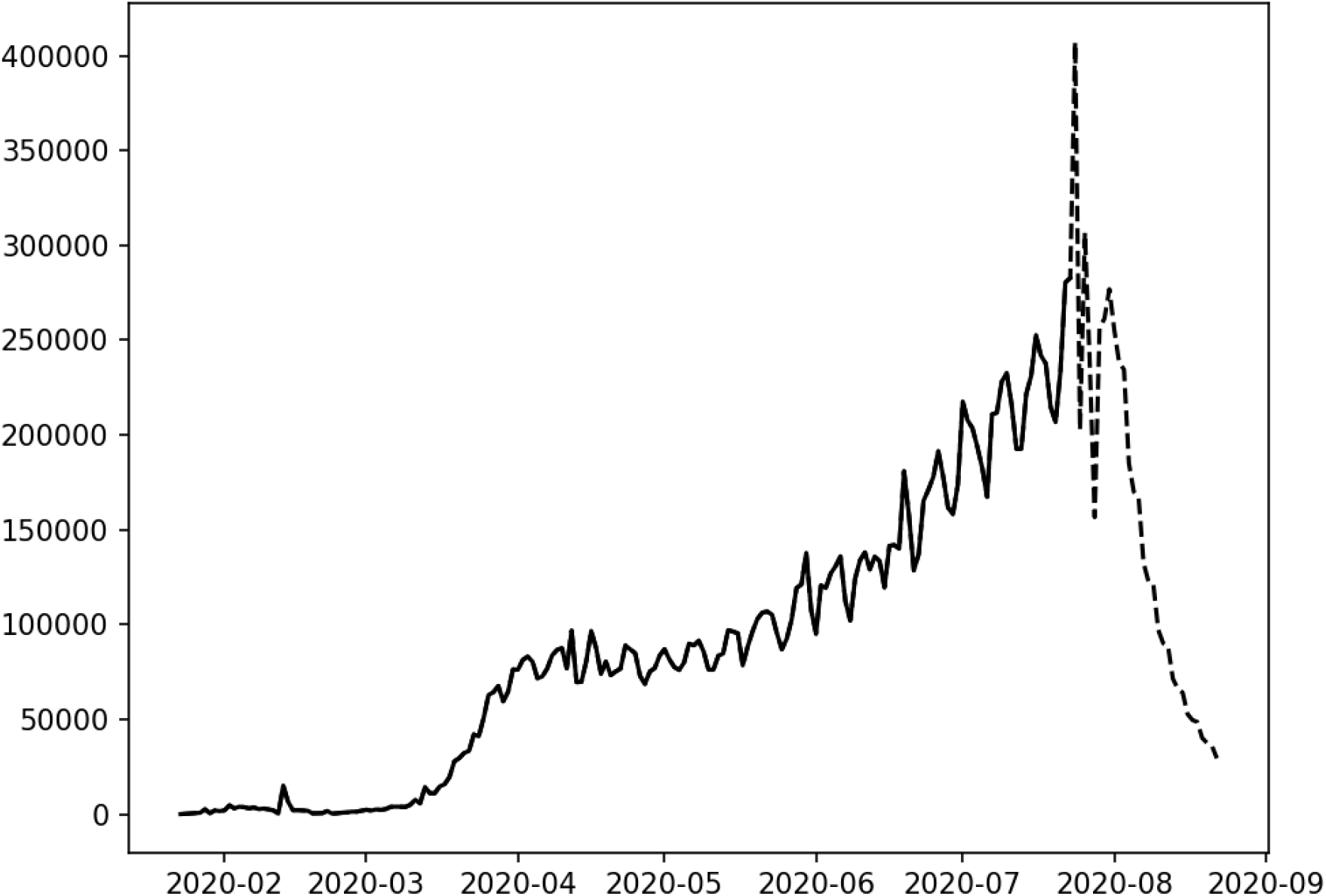
The reported and forecasted curve of number of new cases of Covid-19 worldwide where solid curve and dotted curve were the number of reported and predicted cumulative cases, respectively.

**Figure 8.**
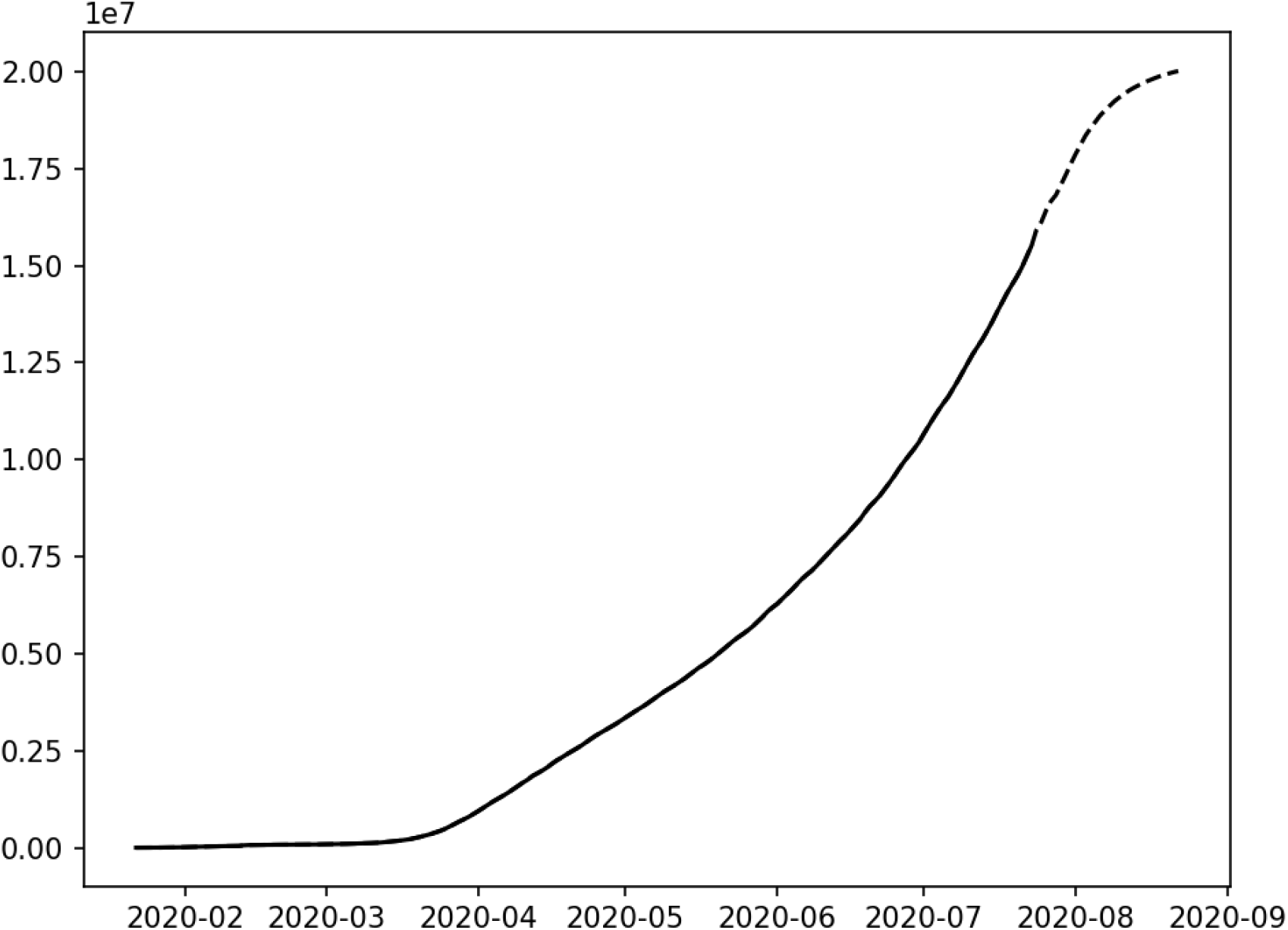
The reported and forecasted curve of number of cumulative cases of Covid-19 worldwide where solid curve and dotted curve were the number of reported and predicted cumulative cases, respectively.

The figures and table demonstrated that the forecasted number of new cases of Covid-19 decreased from 407,205 on July 24, 2020 to 29,517 on August 22, 2020, and the number of cumulative cases of Covid-19 steadily changed from 15,918,430 on July 24 to 20,015,990 cases on August 22. The forecasted number of new cases of Covid-19 has decreased and will continuously decrease for 30 days. The results strongly show that the outbreak of Covid-19 worldwide is curbed.

### Intervention Measure

Traditionally, the effects of the interventions on the transmission dynamics of Covid-19 can be investigated by the reproduction number *R_t_* which measures the average number of individuals one affected individual will transmit the disease to. The reproduction number *R_t_* is often used to determine the dynamic behavior of epidemics. Similar to the reproduction number, we defined an intervention measure *A_t_* to control the spread of Covid-19. Intervention measure was a matric to quantify the degree of control of the intervention action.

Figure 9 plotted the estimated intervention measure curves of the top fifteen most-affected countries as a function of time and Table S3 summarized the estimated intervention measures of the top fifteen most-affected countries. These results showed some patterns of dynamic changes in intervention measures. The shape of the intervention measure curves of these countries characterized the trajectory of Covid-19 in these countries. The common feature of these curves was that both the intervention curve and the number of new case-time curve shared a similar trend. As the number of new cases of Covid-19 increased to peak values, the intervention measure also increased to peak value to curb the growth of the number of new cases. When the number of new cases fluctuated around the peak, the intervention measure also stayed at the plateau for a short time. Then, when the number of new cases decreased toward a small number or zero, the intervention measure decreased and converged to a small stationary value. Intervention measure and the number of new cases of Covid-19 were highly correlated.

To compare the intervention measure *A_t_* with the reproduction number *R_t_*, we downloaded the estimated reproduction number *R_t_* from https://github.com/lin-lab/COVID19-Rt/tree/master/initial estimates, and presented Figure S2 that plotted the reproduction number curves as a function of time in the top fifteen most-affected countries and Figure S3 that plotted both intervention measure *A_t_* and reproduction number *R_t_* curves. In general, the reproduction curves were fluctuated decreasing function except for United Kingdom. When outbreak of Covid-19 began, the reproduction number was in the top of the curve and much larger than 1. As time increased, the reproduction number decreased. When the reproduction number was less than 1, the number of new cases quickly converge to a very small number or to zero. We observed from Figure S3 that the patterns of intervention measure *A_t_* and reproduction number *R_t_* of the top fifteen most-affected countries can be divided into two groups (1) *A_t_* and *R_t_* were positively correlated and (2) *A_t_* and *R_t_* were not correlated or negatively correlated. The first group included US, Spain, Italy, Iran, France, Germany and Turkey and the second group included Brazil, Chile, Russia, India, United Kingdom, Peru, Pakistan, and Mexico. For the countries in the first group, when the reproduction number *R_t_* was large and the spread of Covid-19 was strong, government increased intervention which in turn reduced the reproduction number *R_t_* and then the government relaxed the intervention. As a consequence, the intervention measure *A_t_* and the reproduction number *R_t_* were positively correlated. For the countries in the second group, the health interventions were lag to the reproduction number *R_t_*. When the outbreak of Covid-19 began, the government did not implement strong health interventions, or implement weak interventions, After long time, when the number of virus increased, which in turn, the affected patients were detected and isolated, the reproduction number *R_t_* began to reduce. Due to great pressure from the society, the government’s intervention began to be intensified. The correlation between the intervention measure and the reproduction number showed negative correlation. The second group also included small number of less well controlled countries. The intervention measure in these countries was weak and showed small correlation with the reproduction number.

**Figure 9.**
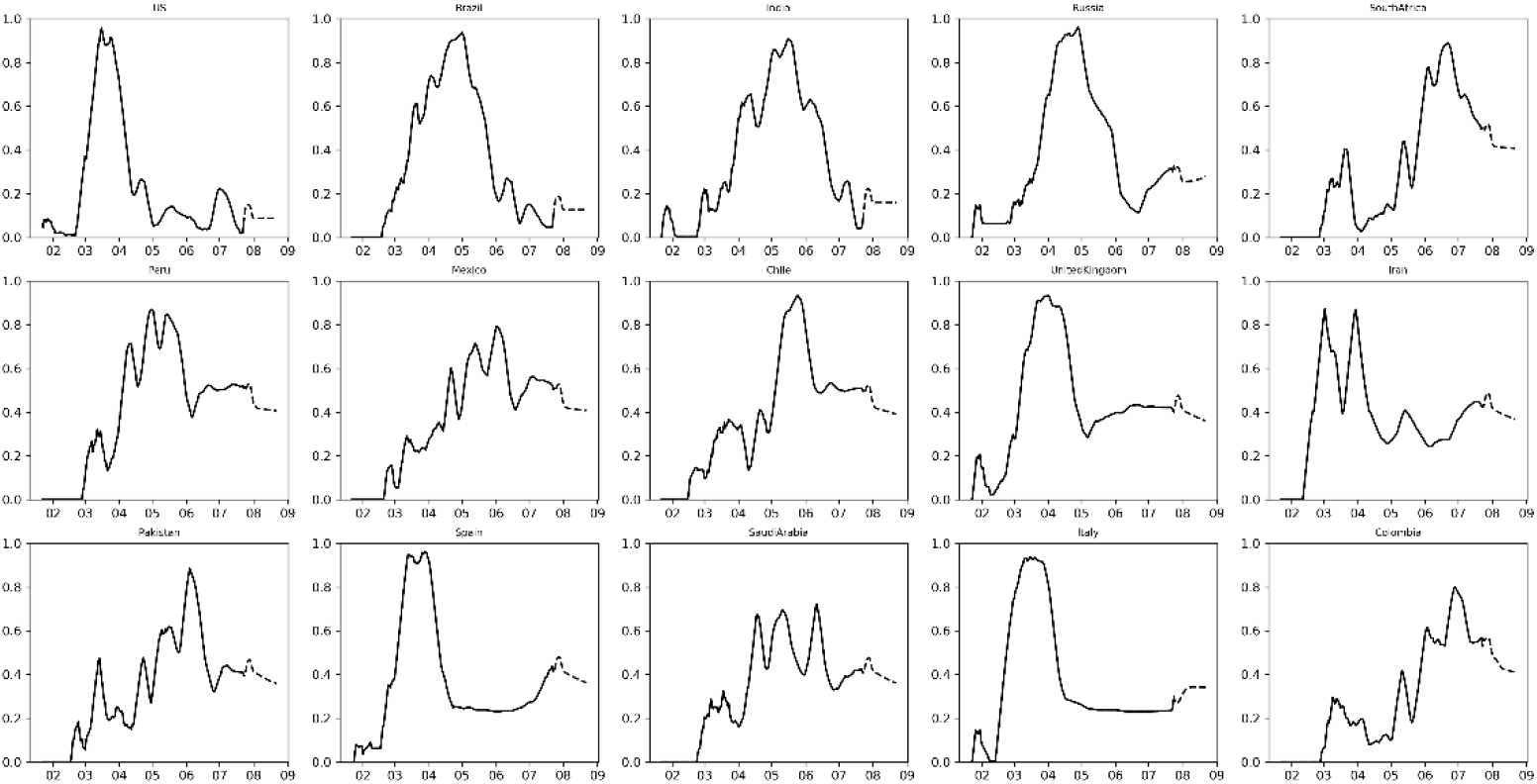
The estimated intervention measures of the top fifteen most-affected countries.

Table S4 summarized the Spearman correlation coefficients between the intervention measures and reproduction number in 187 countries or regions. The Spearman correlation coefficients between the intervention measures and reproduction number in 107 countries or regions were larger than 0.3000, the correlation coefficients in 42 countries or regions were between −0.1000 and 0.3000, and the correlation coefficients in 19 countries and regions were between −0.1000 and −0.5276.

### Clustering Intervention Patterns of the Countries across the World

Clustering algorithm and geographical information system GIS were used to analyze the intervention strategies of all 187 countries across the world. Clustering results would provide information about the spread pattern of the coronavirus across the countries and how to best combat Covid-19. All 187 countries were grouped into 10 clusters using k-means clustering algorithms and intervention measure time curves of the 187 countries across the world (Figure 10 and Table S5).

The first, second and third clusters were the group of the top most-affected countries The first cluster included 9 countries: Brazil, France, Germany, India, Italy, Russia, Spain, Turkey, US. The second cluster included 9 countries: Argentina, Chile, Colombia, Israel, Kyrgyzstan, Mexico, Peru, South Africa. The third cluster included 8 countries: Bangladesh, Iran, Iraq, Pakistan, Qatar, Saudi Arabia, United Kingdom. The shapes of the intervention curves, the production number curves and the number of new case-time curves in these three clusters were the similar. Up to now, the number of cumulative cases of Covid-19 of the countries in these three clusters were the largest.

**Figure 10.**
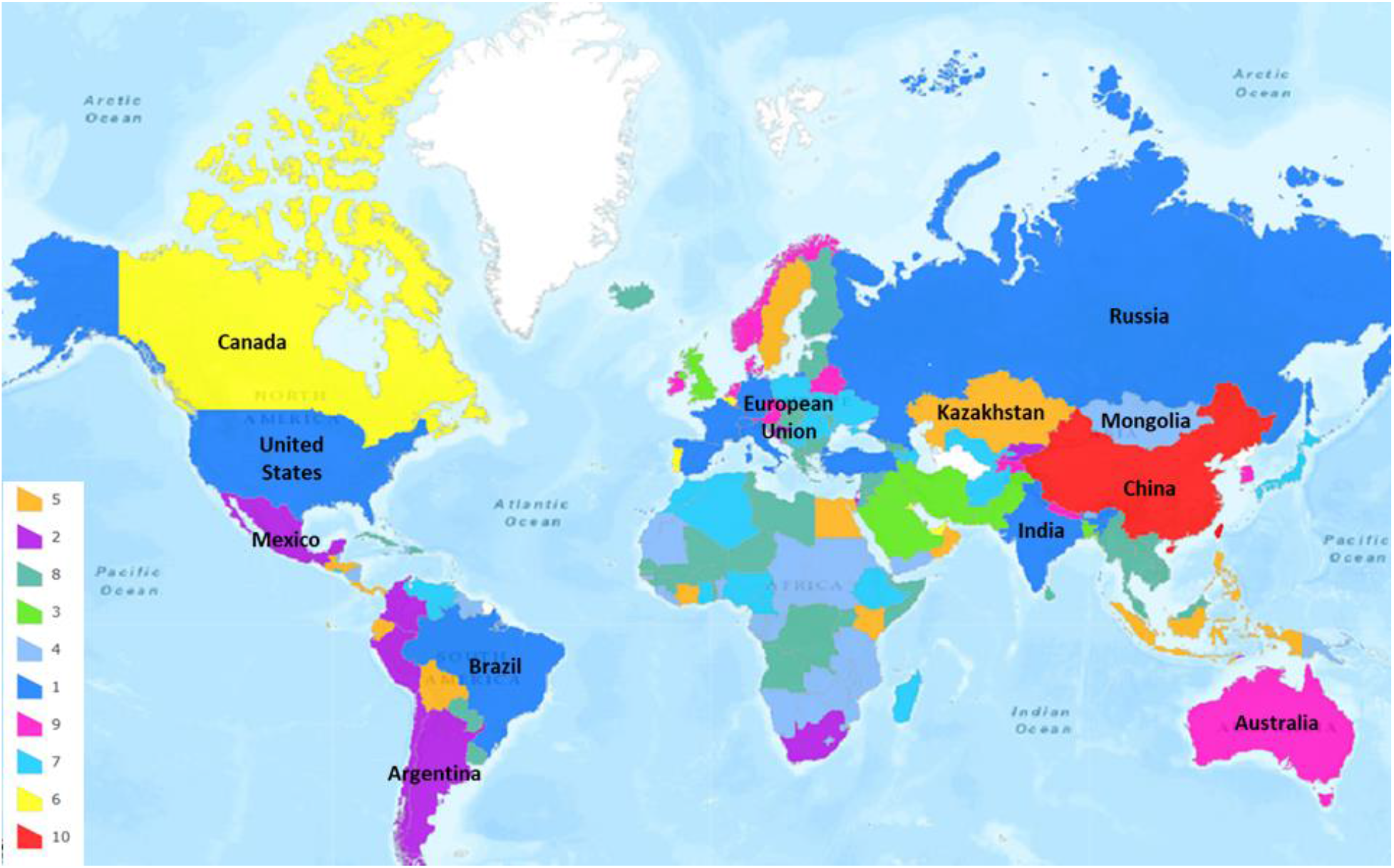
All 187 countries were grouped into ten clusters.

The fifth and sixth lusters were the group of the second most-affected countries. The fifth cluster included 14 countries: Bolivia, Costa Rica, Dominican Republic, Ecuador, Egypt, Guatemala, Honduras, Indonesia, Kazakhstan, Kenya, Oman, Panama, Philippines, Sweden. The sixth cluster included 6 countries: Belgium, Canada, Kuwait, Portugal, Singapore, United Arab Emirates. The countries in these two clusters were the second most-affected countries, but well controlled.

The seventh cluster and ninth cluster were the group of the mildly affected countries. The seventh cluster included 23 countries: Afghanistan, Algeria, Armenia, Azerbaijan, Bahrain, Cameroon, Cote D'ivoire, El Salvador, Ethiopia, Ghana, Japan, Madagascar, Moldova, Montenegro, Morocco, Nigeria, Poland, Romania, Serbia, Ukraine, Uzbekistan, Venezuela, West Bank and Gaza. The ninth cluster included 12 countries: Australia, Austria, Belarus, Czechia, Denmark, Ireland, Korea South, Luxembourg, Nepal, Netherlands, Norway, Tajikistan. The number of cumulative cases of Covid-19 on August 22, 2020 of the countries in the seventh and ninth clusters ranged from 7,187 to 58,865.

The tenth cluster included only China. This was an independent cluster that showed quite different pattern of spread of Covid-19. Countries in the fourth and eighth clusters were less affected.

## Discussion

As an alternative to the epidemiologic transmission models, we developed the RNRL method to help health officers plan public health interventions and combating the spread of Covid-19. We viewed interventions to stop the spread of Covid-19 as actions to control the states of dynamic system and intervention plan as the design of optimal control. A key step for optimal control design was identification of the dynamic system. Therefore, we integrated the identification of the dynamic system underlying Covid-19 and formulated a planning intervention strategy problem as a novel RNRL problem which included recurrent neural network-based reinforcement learning. The RNRL can learn the complex dynamics within the temporal ordering of input time series of Covid-19 and develop suitable interventions for containing the Covid-19.

In this study, we presented a new concept of intervention measure. To improve interpretation of the intervention measure, we compared the intervention measure with the reproduction number. In general, the correlation coefficients between the intervention measure and reproduction number was high except for the less controlled and less affected countries Intervention measure quantified the strength of intervention (control action), while reproduction number measured the state of the spread of Covid-19 being controlled, i.e., measures how well the spread of Covid-19 was curbed. In other words, intervention measure is to quantify how strong the action is, while the reproduction number is to study the effect or the response of intervention. Intervention measure is complimentary to the reproduction number.

The RNRL provided a powerful tool for fighting the surge of Covid-19 worldwide. The dynamic system consists of two essential components. One is the state of the system and the second is action taken. The evolution of the dynamic system highly depends on a sequence of actions. Actions influencing the dynamics of Covid-19 cannot be directly measured or observed. In this report, we proposed to use an intervention measure to quantify the actions. The intervention measure was estimated. The intervention measure curve characterized the dynamics of Covid-19 and can be used to assess the stages of the spread of Covid-19 and strength of the control. The intervention measure curves were used to cluster 187 countries into five basic groups: the most-affected group (26 countries), the second most-affected, but well-control group (20 countries), mildly affected group (35 countries), less affected group (105 countries) and the independent group (China).

Although the number of cumulative cases of Covid-19 worldwide passed 18 million, we are happy to observe that outbreak of Covid-19 worldwide has peaked and is on the decline. The results strongly show that the outbreak of Covid-19 worldwide is curbed. If the less controlled groups of countries continuously strengthen interventions, our analysis demonstrated that the spread of Covid-19 worldwide will be finally stopped. We are confident that we will win the combat to contain the Covid-19.

Since the politics and economics strongly affect the dynamics of Covid-19, the evolutionary trajectories of Covid-19 in most countries will be uncertain. The accuracy of long-term forecasting of Covid-19 may not be very high. However, accuracy of short-term estimation of the number of new cases can be quite good. We suggest that every 10 days we update the data and run the RNRL to forecast the trajectory of Covid-19 in 15 days or one month.

## Data Availability

Data on the number of confirmed and new cases of Covid-19 from January 22, 2020 to June 28, 2020 were obtained from the John Hopkins Coronavirus Resource Center (https://coronavirus.jhu.edu/MAP.HTML).

## Acknowledgements

Dr. Li Jin is partially supported by National Natural Science Foundation of China (91846302). Dr. Wei Lin is supported by the National Key R&D Program of China (Grant no. 2018YFC0116600), the National Natural Science Foundation of China (Grant no. 11925103), and by the STCSM (Grant no. 18DZ1201000).

Many thanks to Ms. Sara A. Barton for editing and the Texas Advanced Computing Center for computation support.

---

**Table S3.** The estimated intervention measures of the top fifteen most-affected countries.

|  | Brazil | Chile |
| --- | --- | --- |
| 2020/1/22 | 0.00000 | 0.00000 |
| 2020/1/23 | 0.00000 | 0.00000 |
| 2020/1/24 | 0.00000 | 0.00000 |
| 2020/1/25 | 0.00000 | 0.00000 |
| 2020/1/26 | 0.00000 | 0.00000 |
| 2020/1/27 | 0.00000 | 0.00000 |
| 2020/1/28 | 0.00000 | 0.00000 |
| 2020/1/29 | 0.00000 | 0.00000 |
| 2020/1/30 | 0.00000 | 0.00000 |
| 2020/1/31 | 0.00000 | 0.00000 |
| 2020/2/1 | 0.00000 | 0.00000 |
| 2020/2/2 | 0.00000 | 0.00000 |
| 2020/2/3 | 0.00000 | 0.00000 |
| 2020/2/4 | 0.00000 | 0.00000 |
| 2020/2/5 | 0.00000 | 0.00000 |
| 2020/2/6 | 0.00000 | 0.00000 |
| 2020/2/7 | 0.00000 | 0.00000 |
| 2020/2/8 | 0.00000 | 0.00000 |
| 2020/2/9 | 0.00000 | 0.00000 |
| 2020/2/10 | 0.00000 | 0.00000 |
| 2020/2/11 | 0.00000 | 0.00000 |
| 2020/2/12 | 0.00000 | 0.00000 |
| 2020/2/13 | 0.00000 | 0.00000 |
| 2020/2/14 | 0.00000 | 0.00000 |
| 2020/2/15 | 0.00000 | 0.00000 |
| 2020/2/16 | 0.00000 | 0.04995 |
| 2020/2/17 | 0.00000 | 0.09801 |
| 2020/2/18 | 0.00000 | 0.10784 |
| 2020/2/19 | 0.04126 | 0.11853 |
| 2020/2/20 | 0.05701 | 0.12921 |

## Supplementary Figure Legend

**Figure S1**. RNRL algorithm flowchart.

**Figure S2**. The reproduction number and intervention measure curves as a function of time in the top fifteen most-affected countries where blue and red curve represented the reproduction number and intervention measure, respectively.

